# Effectiveness of mRNA-1273 against infection and COVID-19 hospitalization with SARS-CoV-2 Omicron subvariants: BA.1, BA.2, BA.2.12.1, BA.4, and BA.5

**DOI:** 10.1101/2022.09.30.22280573

**Authors:** Hung Fu Tseng, Bradley K. Ackerson, Katia J. Bruxvoort, Lina S. Sy, Julia E. Tubert, Gina S. Lee, Jennifer H. Ku, Ana Florea, Yi Luo, Sijia Qiu, Soon Kyu Choi, Harpreet S. Takhar, Michael Aragones, Yamuna D. Paila, Scott Chavers, Carla A. Talarico, Lei Qian

## Abstract

Studies have reported reduced natural SARS-CoV-2 infection- and vaccine-induced neutralization against Omicron BA.4/BA.5 compared with earlier Omicron subvariants. We conducted a test-negative case–control study evaluating mRNA-1273 vaccine effectiveness (VE) against infection and hospitalization with Omicron subvariants. The study included 30,809 SARS-CoV-2 positive and 92,427 SARS-CoV-2 negative individuals aged ≥18 years tested during 1/1/2022-6/30/2022. While 3-dose VE against BA.1 infection was high and waned slowly, VE against BA.2, BA.2.12.1, BA.4, and BA.5 infection was initially moderate to high (61.0%-90.6% 14-30 days post third dose) and waned rapidly. The 4-dose VE against infection with BA.2, BA.2.12.1, and BA.4 ranged between 64.3%-75.7%, and was low (30.8%) against BA.5 14-30 days post fourth dose, disappearing beyond 90 days for all subvariants. The 3-dose VE against hospitalization for BA.1, BA.2, and BA.4/BA.5 was 97.5%, 82.0%, and 72.4%, respectively; 4-dose VE against hospitalization for BA.4/BA.5 was 88.5%. Evaluation of the updated bivalent booster is warranted.

## Main

Since the detection of SARS-CoV-2 in Wuhan, China, in December 2019, several new variants of concern (VOC) have emerged, many of which were associated with pandemic waves^1^. The most recent VOC, Omicron, first detected in South Africa in November 2021, is substantially more transmissible than earlier VOCs^2^ and contains multiple mutations that confer greater escape from naturally acquired and vaccine-elicited immunity compared with earlier variants^3,4^. Together, these characteristics allowed Omicron to rapidly become the dominant strain globally and resulted in large waves of infection much greater than any seen previously during the pandemic^2,5,6^. Within a few months after the emergence of Omicron, the initially dominant subvariant BA.1 was replaced by BA.2 and BA.2.12.1 subvariants, which are more transmissible^5,7,8^ but do not appear to have greater ability to evade vaccine-elicited protection than BA.1^9^. However, soon after, the subvariants BA.4 and BA.5 became the dominant strains globally^10-12^. Several *in vitro* studies reported lower natural SARS-CoV-2 infection- and vaccine-induced neutralization activity against BA.4 and BA.5 than earlier Omicron subvariants, raising concerns about potentially increased escape from natural and vaccine-induced protection^11,13-20^.

Previous studies have shown markedly reduced vaccine effectiveness (VE) of 2 doses of mRNA vaccines, including mRNA-1273 (Spikevax; Moderna Inc, Cambridge, MA, USA) and BNT162b2 (Cominarty; Pfizer Inc, New York, NY, USA; BioNTech Manufacturing GmbH, Mainz, Germany), against infection with BA.1 compared with earlier VOCs^9^. After a third dose, VE of mRNA vaccines initially improved but waned quickly^9,21-25^. We previously found that the 2-dose VE of mRNA-1273 against BA.1 infection was initially 44.0% compared with 80.2% against Delta infection, waning to 5.9% and 61.3% at >270 days, for BA.1 and Delta, respectively^22^. Similarly, 3-dose VE of mRNA-1273 against BA.1 infection decreased from 72.1% to 51.2% at >60 days, while 3-dose VE against Delta infection only declined from 94.2% to 88.1% over the same time interval^22^. Additional studies of mRNA vaccines found that 2-dose VE against hospitalization with BA.1 was modest and waned quickly^23-25^, and while 3-dose VE against hospitalization with BA.1 was initially higher, it also waned^23-28^. Of concern, the substantially greater ability of BA.4 and BA.5 to escape vaccine-elicited immunity compared with BA.1 suggests that there may be even greater declines in VE of current vaccines against the BA.4 and BA.5 subvariants.

Few studies have examined the effectiveness of mRNA vaccines against emerging Omicron subvariants; this research is critical to inform decisions around the need for variant-specific boosters that may offer broader protection against Omicron subvariants. As such, we conducted a test-negative case-control study in the Kaiser Permanente Southern California (KPSC) healthcare system in the United States to evaluate the effectiveness of monovalent mRNA-1273 against infection with and COVID-19 hospitalization for Omicron subvariants, including BA.4 and BA.5.

## Results

We describe the flow of case and control selection in Fig. 1. A total of 123,236 individuals (30,809 test-positive cases and 92,427 test-negative controls) were included in the study. Of the 30,809 cases, 16,418 (53.3%) were successfully sequenced, 93.2% of which had a composite Ct value ≤27, compared to only 13.1% of the failed sequencing samples (Supplementary Table 1). We present the distribution of SARS-CoV-2 variants by mRNA-1273 vaccination status and by month of specimen collection in Supplementary Fig. 1 and Supplementary Fig. 2, respectively. Overall, BA.1 circulated between January and April 2022; BA.2 (excluding BA.2.12.1) appeared at the end of January 2022 and BA.2.12.1 appeared in late March 2022; both subvariants continued to circulate through the remainder of the study period. BA.4 and BA.5 appeared in early May 2022, and the proportion attributed to these subvariants, especially BA.5, rapidly increased in June 2022.

**Table 1.** Characteristics of SARS-CoV-2 test-positive cases and test-negative controls.

|  | **Test Positive** | **Test Negative** | **P** | **ASD** |
| --- | --- | --- | --- | --- |
|  | **N=30809** | **N=92427** |  |  |
| Vaccination status, n (%) |  |  | <0.01 | 0.16 |
| 1 dose | 278 (0.9%) | 877 (0.9%) |  |  |
| 2 dose | 6932 (22.5%) | 18914 (20.5%) |  |  |
| 3 dose | 12724 (41.3%) | 43995 (47.6%) |  |  |
| 4 dose | 966 (3.1%) | 4084 (4.4%) |  |  |
| Unvaccinated | 9909 (32.2%) | 24557 (26.6%) |  |  |
| Age at specimen collection date, years |  |  | 0.19 | 0.01 |
| Mean (sd) | 47.63 (17.07) | 47.76 (17.32) |  |  |
| Median | 46 | 46 |  |  |
| Q1, Q3 | 34, 60 | 34, 61 |  |  |
| Min, max | 18, 105 | 18, 107 |  |  |
| Age at specimen collection date, years, n (%) |  |  | N/A | N/A |
| 18-44 | 14536 (47.2%) | 43608 (47.2%) |  |  |
| 45-64 | 10708 (34.8%) | 32124 (34.8%) |  |  |
| 65-74 | 3271 (10.6%) | 9813 (10.6%) |  |  |
| ≥75 | 2294 (7.4%) | 6882 (7.4%) |  |  |
| Sex, n (%) |  |  | N/A | N/A |
| Female | 17167 (55.7%) | 51501 (55.7%) |  |  |
| Male | 13642 (44.3%) | 40926 (44.3%) |  |  |
| Race/ethnicity, n (%) |  |  | N/A | N/A |
| Non-Hispanic White | 8903 (28.9%) | 26709 (28.9%) |  |  |
| Non-Hispanic Black | 2503 (8.1%) | 7509 (8.1%) |  |  |
| Hispanic | 13869 (45.0%) | 41607 (45.0%) |  |  |
| Non-Hispanic Asian | 3499 (11.4%) | 10497 (11.4%) |  |  |
| Other/Unknown | 2035 (6.6%) | 6105 (6.6%) |  |  |
| Body mass index<sup>a</sup>, kg/m<sup>2</sup>, n (%) |  |  | <0.01 | 0.06 |
| <18.5 | 303 (1.0%) | 987 (1.1%) |  |  |
| 18.5-<25 | 6140 (19.9%) | 18913 (20.5%) |  |  |
| 25-<30 | 8696 (28.2%) | 26804 (29.0%) |  |  |
| 30-<35 | 6351 (20.6%) | 18828 (20.4%) |  |  |
| 35-<40 | 3122 (10.1%) | 9734 (10.5%) |  |  |
| 40-<45 | 1366 (4.4%) | 4253 (4.6%) |  |  |
| ≥45 | 933 (3.0%) | 2857 (3.1%) |  |  |
| Unknown | 3898 (12.7%) | 10051 (10.9%) |  |  |
| Smoking<sup>a</sup>, n (%) |  |  | <0.01 | 0.06 |
| No | 22543 (73.2%) | 67110 (72.6%) |  |  |
| Yes | 5206 (16.9%) | 17359 (18.8%) |  |  |
| Unknown | 3060 (9.9%) | 7958 (8.6%) |  |  |
| Charlson comorbidity index score<sup>b</sup>, n (%) |  |  | <0.01 | 0.07 |
| 0 | 21590 (70.1%) | 61993 (67.1%) |  |  |
| 1 | 4442 (14.4%) | 13821 (15.0%) |  |  |
| ≥2 | 4777 (15.5%) | 16613 (18.0%) |  |  |
| Frailty index<sup>b</sup> |  |  | <0.01 | 0.06 |
| Mean (sd) | 0.12 (0.03) | 0.12 (0.03) |  |  |
| Median | 0.11 | 0.11 |  |  |
| Q1, Q3 | 0.10, 0.13 | 0.10, 0.13 |  |  |
| Min, max | 0.05, 0.40 | 0.05, 0.42 |  |  |
| Frailty index<sup>b</sup>, n (%) |  |  | <0.01 | 0.08 |
| Quartile 1 | 7169 (23.3%) | 21404 (23.2%) |  |  |
| Quartile 2 | 8926 (29.0%) | 24117 (26.1%) |  |  |
| Quartile 3 | 7666 (24.9%) | 23147 (25.0%) |  |  |
| Quartile 4, most frail | 7048 (22.9%) | 23759 (25.7%) |  |  |
| Chronic diseases<sup>b</sup>, n (%) |  |  |  |  |
| Kidney disease | 1623 (5.3%) | 5549 (6.0%) | <0.01 | 0.03 |
| Heart disease | 1069 (3.5%) | 3632 (3.9%) | <0.01 | 0.02 |
| Lung disease | 2842 (9.2%) | 9495 (10.3%) | <0.01 | 0.04 |
| Liver disease | 1155 (3.7%) | 4137 (4.5%) | <0.01 | 0.04 |
| Diabetes | 4313 (14.0%) | 13766 (14.9%) | <0.01 | 0.03 |
| Immunocompromised, n (%) | 1165 (3.8%) | 4326 (4.7%) | <0.01 | 0.04 |
| HIV/AIDS, n | 78 | 365 |  |  |
| Leukemia/lymphoma, congenital and other immunodeficiencies, asplenia/hyposplenia, n | 425 | 1443 |  |  |
| Hematopoietic stem cell transplant/organ transplant, n | 145 | 459 |  |  |
| Immunosuppressant medications, n | 785 | 2906 |  |  |
| Autoimmune conditions<sup>b</sup>, n (%) | 922 (3.0%) | 3110 (3.4%) | <0.01 | 0.02 |
| Rheumatoid arthritis, n | 388 | 1340 |  |  |
| Inflammatory bowel disease, n | 194 | 705 |  |  |
| Psoriasis and psoriatic arthritis, n | 326 | 973 |  |  |
| Multiple sclerosis, n | 54 | 165 |  |  |
| Systemic lupus erythematosus, n | 79 | 317 |  |  |
| Pregnant at specimen collection date, n (%) | 742 (2.4%) | 3954 (4.3%) | <0.01 | 0.10 |
| 1st trimester, n | 126 | 370 |  |  |
| 2nd trimester, n | 216 | 533 |  |  |
| 3rd trimester, n | 400 | 3051 |  |  |
| History of SARS-CoV-2 infection<sup>c</sup>, n (%) | 4512 (14.6%) | 19811 (21.4%) | <0.01 | 0.18 |
| History of SARS-CoV-2 molecular test<sup>c</sup>, n (%) | 23802 (77.3%) | 64378 (69.7%) | <0.01 | 0.17 |
| Number of outpatient and virtual visits<sup>b</sup>, n (%) |  |  | <0.01 | 0.15 |
| 0 | 2157 (7.0%) | 5649 (6.1%) |  |  |
| 1-4 | 8192 (26.6%) | 19958 (21.6%) |  |  |
| 5-10 | 8951 (29.1%) | 26395 (28.6%) |  |  |
| ≥11 | 11509 (37.4%) | 40425 (43.7%) |  |  |
| Number of emergency department visits<sup>b</sup>, n (%) |  |  | <0.01 | 0.07 |
| 0 | 24801 (80.5%) | 71843 (77.7%) |  |  |
| 1 | 3920 (12.7%) | 13648 (14.8%) |  |  |
| ≥2 | 2088 (6.8%) | 6936 (7.5%) |  |  |
| Number of hospitalizations<sup>b</sup>, n (%) |  |  | <0.01 | 0.03 |
| 0 | 29016 (94.2%) | 86543 (93.6%) |  |  |
| 1 | 1391 (4.5%) | 4437 (4.8%) |  |  |
| ≥2 | 402 (1.3%) | 1447 (1.6%) |  |  |
| Preventive care<sup>b</sup>, n (%) | 21899 (71.1%) | 69064 (74.7%) | <0.01 | 0.08 |
| Medicaid, n (%) | 2941 (9.5%) | 10057 (10.9%) | <0.01 | 0.04 |
| Neighborhood median household income, n (%) |  |  | 0.02 | 0.02 |
| <\$40,000 | 1099 (3.6%) | 3545 (3.8%) | | |
| \$40,000-\$59,999 | 5657 (18.4%) | 17505 (18.9%) | | |
| \$60,000-\$79,999 | 7710 (25.0%) | 22652 (24.5%) | | |
| \$80,000+ | 16324 (53.0%) | 48665 (52.7%) | | |
| Unknown | 19 (0.1%) | 60 (0.1%) |  |  |
| KPSC physician/employee, n (%) | 1755 (5.7%) | 3664 (4.0%) | <0.01 | 0.08 |
| Medical center area<sup>d</sup> |  |  | <0.01 | 0.15 |
| Month of specimen collection, n (%) |  |  | <0.01 | 0.23 |
| January 2022 | 6797 (22.1%) | 23700 (25.6%) |  |  |
| February 2022 | 4602 (14.9%) | 9108 (9.9%) |  |  |
| March 2022 | 1311 (4.3%) | 6199 (6.7%) |  |  |
| April 2022 | 2613 (8.5%) | 11177 (12.1%) |  |  |
| May 2022 | 7828 (25.4%) | 21047 (22.8%) |  |  |
| June 2022 | 7658 (24.9%) | 21196 (22.9%) |  |  |
| Specimen type, n (%) |  |  | 0.85 | <0.01 |
| Nasopharyngeal/oropharyngeal swab | 25387 (82.4%) | 76116 (82.4%) |  |  |
| Saliva | 5422 (17.6%) | 16311 (17.6%) |  |  |

<sup>a</sup> Defined in the 2 years prior to specimen collection date.
<sup>b</sup> Defined in the 1 year prior to specimen collection date.
<sup>c</sup> Defined based on all available medical records from March 1, 2020, to specimen collection date.
<sup>d</sup> Frequency and percent for the 19 medical center areas not shown.
N/A, not applicable.

**Fig. 1:**
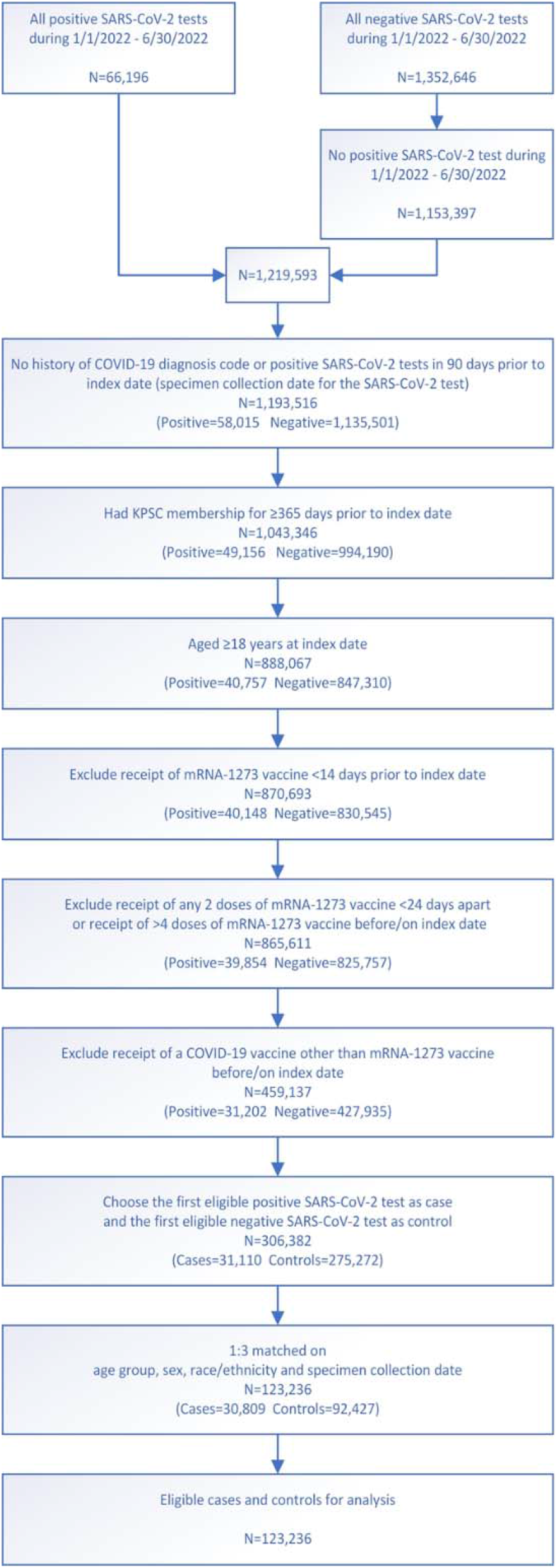
Flow chart for mRNA-1273 vaccine test-negative design.

**Fig. 2:**
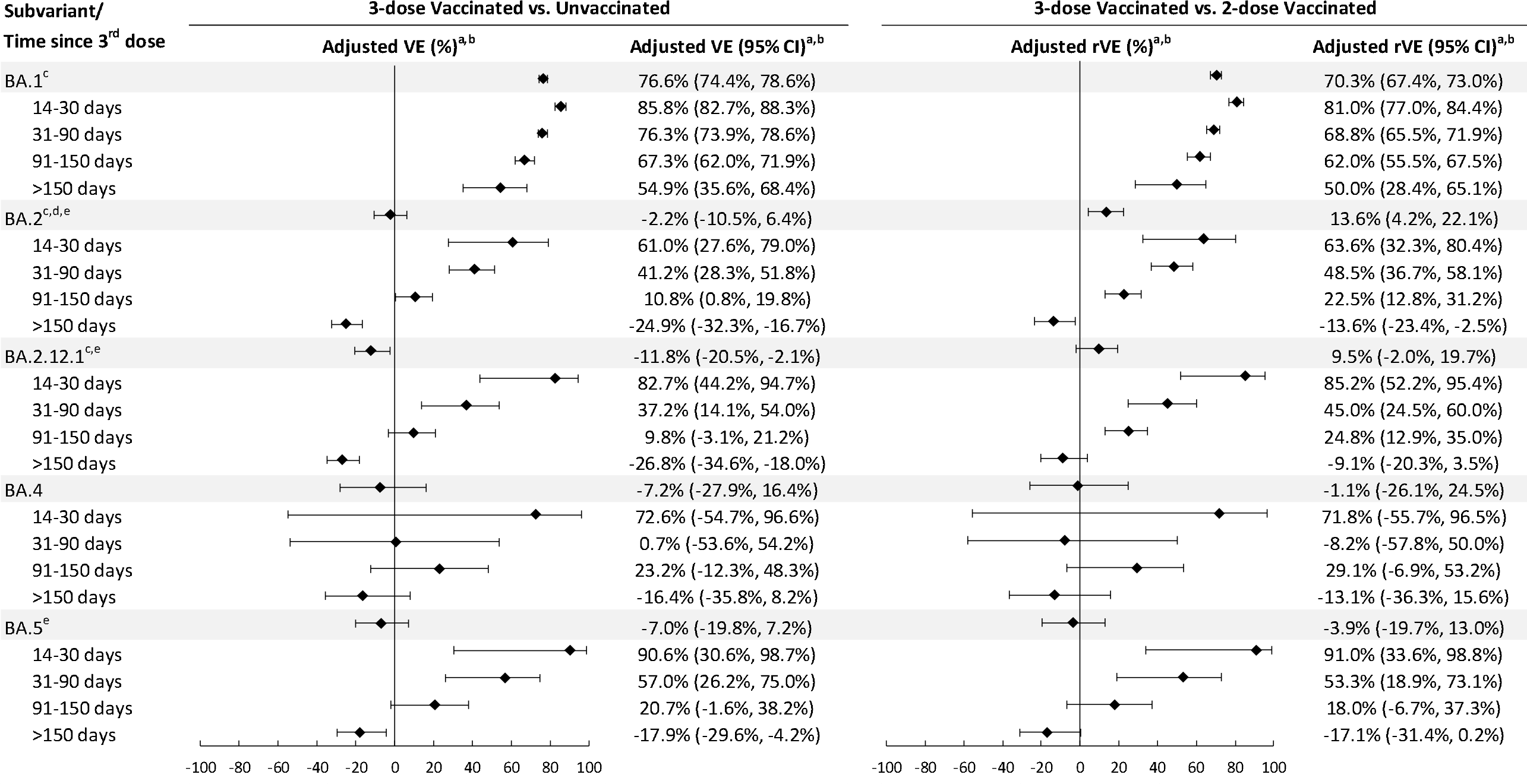
Vaccine effectiveness of 3 doses of mRNA-1273 and relative vaccine effectiveness of 3 versus 2 doses of mRNA-1273 against infection with SARS-CoV-2 variants by time since vaccination. ^a^When the OR or its 95% CI was >1, the VE/rVE or its 95% CI was transformed as ([1/OR] – 1) x 100. ^b^ Adjusted for age, sex, race/ethnicity, month of specimen collection, history of SARS-CoV-2 infection, history of SARS-CoV-2 molecular test, number of outpatient and virtual visits, medical center area, and time between second dose and specimen collection date (for 3-dose versus 2-dose models only). ^c^ Medical center area removed from adjustment set in 3-dose versus 2-dose models due to lack of model convergence. ^d^BA.2 excluding BA.2.12.1. ^e^Medical center area removed from adjustment set in 3-dose versus unvaccinated models due to lack of model convergence. CI, confidence interval; OR, odds ratio; rVE, relative vaccine effectiveness; VE, vaccine effectiveness.

We described baseline characteristics of cases and controls in Table 1. The median age of individuals included in the study was 46 years, of which 18% were aged ≥65 years. Females accounted for 55.7% of the total population. Forty-five percent of the individuals were Hispanic, 28.9% non-Hispanic White, 8.1% non-Hispanic Black, 11.4% non-Hispanic Asian, and 6.6% other or unknown race/ethnicity. Cases and controls had similar distribution of many covariates (absolute standardized difference [ASD]≤0.1), including body mass index, smoking history, Charlson comorbidity score, frailty index, prevalence of chronic diseases, pregnancy status, immunocompromised status, prevalence of autoimmune conditions, history of emergency department (ED) visits, history of hospitalization, use of preventive care, Medicaid status, neighbourhood median household income, KSPC physician/employee status, and specimen type.

In analyses of 3-dose VE (versus unvaccinated) against infection with Omicron subvariants by time since vaccination, the 3-dose VE against BA.1 ranged from 85.8% (95% confidence interval [CI] 82.7%, 88.3%) in the 14-30 days after the third dose to 54.9% (95% CI 35.6%, 68.4%) >150 days after the third dose (Fig. 2, Supplementary Table 2a). VE for these two time intervals, respectively, was 61.0% (95% CI 27.6%, 79.0%) and -24.9% (95% CI -32.3%, -16.7%) for BA.2, excluding BA.2.12.1; 82.7% (95% CI 44.2%, 94.7%) and -26.8% (95% CI -34.6%, -18.0%) for BA.2.12.1; 72.6% (95% CI - 54.7%, 96.6%) and -16.4% (95% CI -35.8%, 8.2%) for BA.4; and 90.6% (95% CI 30.6%, 98.7%) and - 17.9% (95% CI -29.6%, -4.2%) for BA.5. We also present the relative VE (rVE) comparing 3 doses to 2 doses against Omicron subvariants by time since vaccination (Fig. 2, Supplementary Table 2b). In general, we observed consistent incremental protection of 3 doses versus 2 doses in the 14-90 days after the third dose, other than against BA.4, which had a small number of cases and wide CI. The incremental benefit in protection decreased by time since the third dose. For BA.5, the 95% CI of rVE included 0 after >90 days after the third dose.

In analyses of 4-dose VE against infection with Omicron subvariants by time since vaccination, because a fourth dose (second booster) in adults ages ≥50 years was not recommended until the tail-end of the BA.1 period, there were insufficient numbers to estimate 4-dose VE against BA.1. The 4-dose VE against BA.2 was 64.3% (95% CI 50.7%, 74.2%) 14-30 days after the fourth dose and 17.3% (95% CI -45.3%, 62.6%) >90 days after the fourth dose (Fig. 3, Supplementary Table 3a). VE for these time intervals, respectively, was 64.4% (95% CI 48.6%, 75.4%) and 14.0% (95% CI -48.4%, 61.9%) for BA.2.12.1; 75.7% (95% CI 34.7%, 91.0%) and 6.3% (95% CI -66.3%, 70.4%) for BA.4; and 30.8% (95% CI -9.2%, 56.5%) and 5.0% (95% CI -56.9%, 61.1%) for BA.5. We also present the rVE comparing 4 doses to 3 doses against Omicron subvariants by time since vaccination (Fig. 3, Supplementary Table 3b). We observed consistent incremental protection of 4 doses compared with 3 doses in the 14-90 days after the fourth dose. For BA.4 and BA.5, the 95% CI of rVE included 0 after >90 days after the fourth dose.

**Fig. 3:**
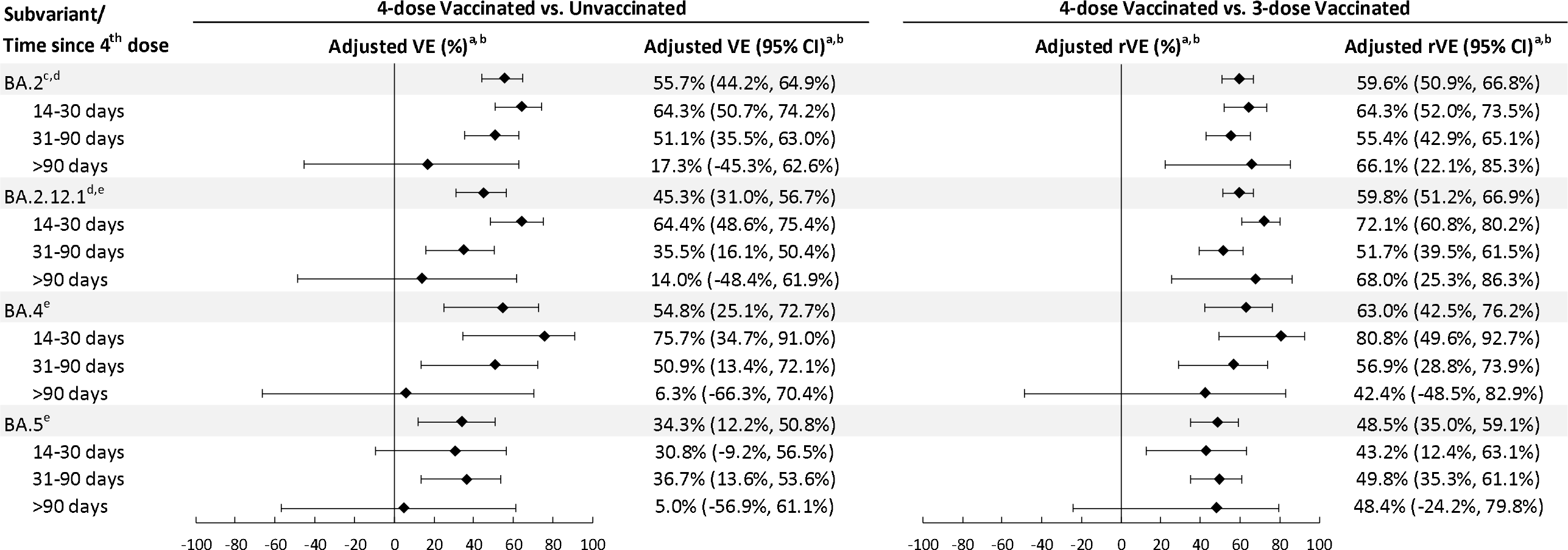
Vaccine effectiveness of 4 doses of mRNA-1273 and relative vaccine effectiveness of 4 versus 3 doses of mRNA-1273 against infection with SARS-CoV-2 variants by time since vaccination. ^a^When the OR or its 95% CI was >1, the VE/rVE or its 95% CI was transformed as ([1/OR] – 1) x 100. ^b^Adjusted for age, sex, race/ethnicity, month of specimen collection, history of SARS-CoV-2 infection, history of SARS-CoV-2 molecular test, number of outpatient and virtual visits, medical center area, and time between third dose and specimen collection date (for 4-dose versus 3-dose models only). ^c^BA.2 excluding BA.2.12.1. ^d^Medical center area removed from adjustment set in 4-dose versus 3-dose models due to lack of model convergence. ^e^Medical center area removed from adjustment set in 4-dose versus unvaccinated models due to lack of model convergence. CI, confidence interval; OR, odds ratio; rVE, relative vaccine effectiveness; VE, vaccine effectiveness.

We examined 3-dose VE against COVID-19 hospitalization for BA.1, BA.2 (including BA.2.12.1), and BA.4/BA.5. The 3-dose VE against hospitalization for BA.1 was 97.5% (95% CI 96.3%, 98.3%) (Fig. 4, Supplementary Table 4a). The 3-dose VE against hospitalization for BA.2 was 82.0% (95% CI 64.5%, 90.8%), while 3-dose VE against hospitalization for BA.4/BA.5 was 72.4% (95% CI 23.9%, 90.0%). We also present the rVE comparing 3 doses to 2 doses against hospitalization for BA.1, BA.2, and BA.4/BA.5 (Fig. 4, Supplementary Table 4b). The rVE against these Omicron subvariants, respectively, was 88.8% (95% CI 83.3%, 92.5%), 75.0% (95% CI 47.6%, 88.1%), and 87.5% (95% CI 51.8%, 96.8%).

**Fig. 4:**
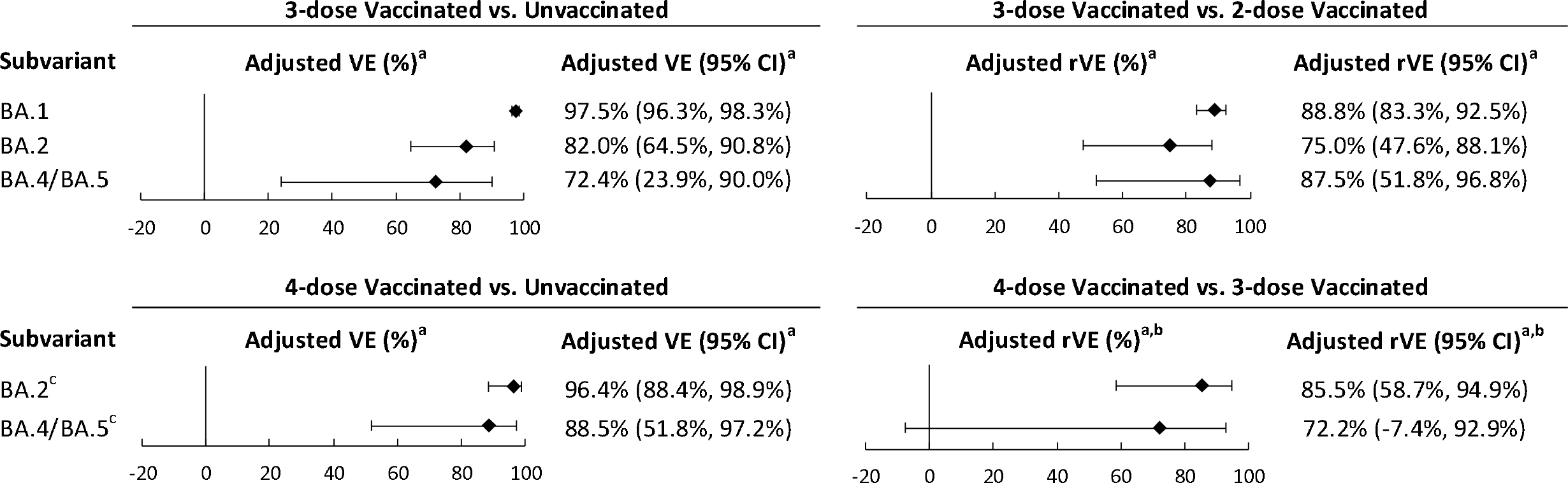
Vaccine effectiveness of 3 and 4 doses of mRNA-1273 and relative vaccine effectiveness of 3 versus 2 doses and 4 versus 3 doses of mRNA-1273 against COVID-19 hospitalization associated with SARS-CoV-2 variants. ^a^Adjusted for age, sex, race/ethnicity, month of specimen collection, history of SARS-CoV-2 infection, history of SARS-CoV-2 molecular test, number of outpatient and virtual visits, and time between second/third dose and specimen collection date (for 3-dose versus 2-dose and 4-dose versus 3-dose models, respectively). Medical center area dropped from adjustment set due to lack of model convergence. ^b^When the OR or its 95% CI was >1, the VE/rVE or its 95% CI was transformed as ([1/OR] – 1) x 100. ^c^History of SARS-CoV-2 infection dropped from adjustment set in 4-dose versus 3-dose models due to lack of model convergence. CI, confidence interval; OR, odds ratio; rVE, relative vaccine effectiveness; VE, vaccine effectiveness.

In the analyses of 4-dose VE against COVID-19 hospitalization for BA.2 (including BA.2.12.1) and BA.4/BA.5, the 4-dose VE against hospitalization for BA.2 was 96.4% (95% CI 88.4%, 98.9%) (Fig. 4, Supplementary Table 5a). The 4-dose VE against hospitalization for BA.4/BA.5 was 88.5% (95% CI 51.8%, 97.2%). We also present the rVE comparing 4 doses to 3 doses against hospitalization for BA.2 and BA.4/BA.5 (Fig. 4, Supplementary Table 5b). The rVE against these Omicron subvariants, respectively, was 85.5% (95% CI 58.7%, 94.9%) and 72.2% (95% CI -7.4%, 92.9%).

In the sensitivity analyses, we imputed Omicron subvariant information using available S-gene target failure (SGTF) data for 12,006 specimens (83.4%) that were not successfully sequenced. By comparing whole genome sequencing (WGS) results and available SGTF results, the positive predictive value of using SGTF results combined with calendar month to predict Omicron subvariants BA.1, BA.2, and BA.4/BA.5 was 99.9%, 99.1%, and 96.1%, respectively. The VE and rVE results are presented in Supplementary Tables 6a-9b. In general, the increased sample size allowed for more precise estimation of VE and rVE against infection and hospitalization, as shown in narrower CIs. The VE and rVE against infection generally appeared to be lower than VE and rVE estimates that included successfully sequenced samples only. The VE and rVE against hospitalization were less impacted in the sensitivity analyses because imputation of SGTF data did not substantively change the numbers.

In the sensitivity analysis excluding immunocompromised individuals, the VE or rVE estimates against infection generally did not vary substantially from the main analyses, except that the point estimates of VE against BA.2 and BA.5 infection at >90 days after the fourth dose were higher; however, the CI still included 0. This also translated to higher rVE of 4 doses compared with 3 doses against the two subvariants, compared with the main analyses. In addition, the point estimate for 3-dose VE against hospitalization for BA.4/BA.5 was higher in the sensitivity analysis (88.5% [95% CI 61.0%, 96.6%] compared with 72.4% (95% CI 23.9%, 90.0%) in the main analysis (Supplementary Tables 10a-13b).

## Discussion

The study evaluated the effectiveness of 3 and 4 doses of mRNA-1273 against infection with and hospitalization for Omicron subvariants in a large, racially, ethnically, and socioeconomically diverse population. The rapid emergence of several subvariants of Omicron, particularly BA.4 and BA.5, which have markedly increased transmissibility and ability to evade natural and vaccine-elicited immunity, raise concerns about the ability of original monovalent COVID-19 vaccines to protect against SARS-COV-2 infections^2,16^. Using successfully sequenced results, we were able to focus our analyses on cases that tended to have a higher viral load and were more likely symptomatic. In addition, COVID-19 hospitalized cases met a prespecified, previously validated case definition or charts were reviewed to confirm hospitalization for severe COVID-19, rather than hospitalization that was coincident with COVID-19. The results provide relevant evidence of mRNA-1273 effectiveness in alleviating COVID-related disease burden in a real-world setting.

Our study found that 3-dose VE of mRNA-1273 against infection with BA.1 was high and waned slowly, whereas VE against infection with more recent Omicron subvariants, including BA.2, BA.2.12.1, BA.4, and BA.5, waned more rapidly. Similarly, 4-dose VE against infection with BA.2, BA.2.12.1, BA.4 and BA.5 was moderate, and was only approximately 35% against BA.5. The 4-dose VE against these subvariants was short-lived, disappearing beyond 90 days after the fourth dose. In a recent study, Qu et al. indicated that although the decay rate of booster neutralizing antibody was similar among variants, the Omicron subvariants, especially BA.4 and BA.5, had substantial neutralization resistance. Their data suggest that both SARS-CoV-2 variant evolution and waning neutralizing antibody titers reduce booster-induced immune protection^29^. Our results also suggest that monovalent Wuhan strain formulated mRNA vaccine would be inadequate in controlling a potential BA.5 surge during the winter season.

Although 3-dose VE of mRNA-1273 against hospitalization was much higher than VE against infection for all Omicron subvariants assessed, the 3-dose VE against hospitalization for BA.2 and especially against hospitalization for BA.4/BA.5 was lower than that against hospitalization for BA.1. The results from sensitivity analyses suggest the 3-dose VE against hospitalization for BA.4/BA.5 could be particularly low for immunocompromised individuals. Compared with 3 doses of mRNA-1273, 4 doses confer additional protection against hospitalization for either BA.2 or BA.4/BA.5. The durability against hospitalization for BA.4/BA.5 is still unknown. Monitoring for waning protection against hospitalization for BA.4/BA.5 or subsequent new subvariants that may emerge will be critically important as more data becomes available^19^.

While prior data on the effectiveness of mRNA-1273 against infection with or hospitalization for BA.4/BA.5 is limited, Hansen et al found that 3 doses of mRNA vaccine provided similar protection against infection with BA.2 or BA.5. However, there was a trend towards increased risk of infection with BA.5 compared with BA.2 among those who had received a second dose more than 4.5 months prior to the third dose^30^. In addition, we and others have previously demonstrated markedly reduced 3-dose mRNA-1273 VE against infection with BA.1 that waned quickly compared to VE that had been observed against earlier variants^9,22^. Similar findings were seen for VE of BNT162b2 or mRNA-based vaccines combined against infection or ED and urgent care encounters^21,23,24^. A fourth mRNA vaccine dose was found to provide some additional short-term protection against infection or ED and urgent care encounters with BA.1 that waned quickly^24,31-35^. However, while short-term 3-dose mRNA VE against severe outcomes with BA.1 was initially found to be high, similar to our findings, it was lower against BA.2 than BA.1 and waned quickly^9,22-24,26^. However, protection from a fourth dose of mRNA vaccines against severe disease with BA.1 was found to be greater and more durable^32,33^. These data suggest that Omicron-specific boosters are likely needed to maintain protection against Omicron subvariants and minimize waning effectiveness.

This study provides important data on the effectiveness of mRNA-1273 against infection with and hospitalization for Omicron subvariants, including predominant subvariants, BA.4 and BA.5. This study has several strengths and limitations. First, the results of our test-negative case-control study may not be generalizable to people who are not tested for SARS-CoV-2, including those with milder symptoms who may not seek testing in healthcare settings. There are several risk factors for infections or severe outcomes that may be associated with both testing and vaccination that could introduce bias, for example, mask use, social distancing, and hygiene practices. We attempted to reduce potential bias by adjusting for sociodemographic characteristics, prior healthcare use, prior SARS-CoV-2 testing, and comorbidities in the models, but residual confounding may remain. For example, some negative VE estimates observed at >150 days after vaccination could be due to differential risk behaviors among vaccinated and unvaccinated individuals when protection from antibodies becomes minimal. Second, as predominant subvariants evolved during the study period, many other factors could also change over time, such as practice of non-pharmacologic interventions, availability of antiviral medications or monoclonal antibody treatments, preventive public health policy, and individual behaviors. These changes might impact the comparison of VE across subvariants.

Third, misclassification of test-positive cases and test-negative controls was another possible source of bias. However, we used a highly specific and sensitive RT-PCR test that minimized misclassification and allowed us to monitor variant proportions through WGS and SGTF analysis. Similarly, misclassification of vaccination status was possible but likely minimal, as KPSC electronic health records (EHRs) captured all vaccinations administered within KPSC and were updated daily with vaccine administration data from the California Immunization Registry, to which all facilities are required by law to report COVID-19 vaccinations within 24 hours. In addition, inclusion of patients hospitalized for reasons other than COVID-19 who are found to have coincident SARS-CoV-2 infection with minimal or no symptoms could also introduce bias^36-38^. In this study, hospitalizations for COVID-19 were identified using a prespecified algorithm, or charts were reviewed to confirm severe COVID-19 disease leading to hospitalization, decreasing the possibility of spuriously reduced estimates of VE against severe disease^38^.

Fourth, statistical power might have been insufficient for testing VE against some subvariants that had lower numbers of cases, resulting in wide confidence intervals for some VE estimates. This was addressed by the sensitivity analysis using SGTF results, in which the VE estimates became more precise. Finally, multiple comparisons were not adjusted for in the analyses, as the focus of the study was on estimating clinically meaningful VE over time across subvariants, rather than statistical significance.

In conclusion, our data indicate that the 3-dose or 4-dose effectiveness of mRNA-1273 against infection with Omicron subvariants is moderate and short-lived, but protection against severe COVID-19 disease remains robust. With the updated bivalent BA.4/BA.5–containing booster (mRNA-1273.222) available in the United States, it is imperative to continue to evaluate its effectiveness, durability, and impact on SARS-CoV-2 evolution.

## Methods

### Study setting

KPSC is an integrated health system that provides healthcare services and insurance coverage to more than 4.7 million members with sociodemographic characteristics representative of the diverse Southern California population^39,40^. EHRs comprehensively capture details of patient care, including vaccinations, diagnoses, laboratory tests, procedures, and pharmacy records. Although most members seek care at KPSC facilities (i.e., 15 hospitals and 236 medical offices), care received outside of KPSC is incorporated into the EHR as part of claims reimbursement. In addition, vaccinations received outside of KPSC are imported daily from external sources, including the California Immunizations Registry (CAIR), Care Everywhere (system on the Epic EHR platform that allows different healthcare systems to exchange patients’ medical information), claims (for example, retail pharmacies), and self-report by members (with valid documentation). The study was approved by the KPSC Institutional Review Board, which waived requirements for written informed consent and written Health Insurance Portability and Accountability Act authorization, as the use of EHRs for this observational study involved minimal risk.

### Laboratory methods

SARS-CoV-2 molecular diagnostic testing is conducted routinely at KPSC for members with and without symptoms who request testing for any reason and prior to certain procedures or hospital admission. Nasopharyngeal specimens (for symptomatic or asymptomatic individuals) or saliva specimens (for asymptomatic individuals) are tested using the RT-PCR TaqPath COVID-19 High-Throughput Combo Kit (Thermo Fisher Scientific, CA, USA). SGTF is defined as a SARS-CoV-2 positive specimen with N and ORF1ab genes detected (cycle threshold values <37), but with undetected S gene. Random samples of SARS-CoV-2–positive specimens are sent on a weekly basis to a commercial laboratory for WGS, as detailed in our prior publications^22,40^.

### Study design

We used a test-negative case-control design to assess the effectiveness of 3 and 4 doses of mRNA-1273 against SARS-CoV-2 Omicron subvariants. Cases were identified from individuals with positive SARS-CoV-2 RT-PCR tests from specimens collected between 1/1/2022 and 6/30/2022 that were sent for WGS and controls that were identified from those with only negative SARS-CoV-2 RT-PCR tests during the same period. Individuals were included if they were aged ≥18 years and had ≥12 months of KPSC membership before the specimen collection date (necessary for accurate ascertainment of exposure status and covariates) and were excluded if they had a history of SARS-CoV-2 infection in the 90 days prior to specimen collection date, received any dose of mRNA-1273 <14 days before the specimen collection date, received any 2 doses of mRNA-1273 <24 days apart or >4 doses of mRNA-1273 before the specimen collection date, or received a COVID-19 vaccine other than mRNA-1273. The first eligible positive and negative SARS-CoV-2 RT-PCR tests were included. We matched cases and controls by a ratio of 1 to 3 on age (18–44 years, 45–64 years, 65–74 years, and ≥75 years), sex, race/ethnicity (non-Hispanic White, non-Hispanic Black, Hispanic, non-Hispanic Asian, and Other/Unknown), and specimen collection date (±10 days).

### Outcomes

Cases consisted of persons infected with BA.1, BA.2 (excluding BA.2.12.1), BA.2.12.1, BA.4, or BA.5, the Omicron subvariants monitored by the World Health Organization that were circulating during the study period^6^. Other variants (e.g., Delta, BA.2.75, BA.3, and recombinant lineages) were not analyzed due to low prevalence during the study period. COVID-19 hospitalization was defined as hospitalization *for* severe COVID-19, rather than hospitalization *with* coincident SARS-CoV-2 infection^36^. COVID-19 hospitalization was initially identified as a SARS-CoV-2–positive test ≤7 days prior to or during hospitalization and further confirmed by (1) ≥1 documented oxygen saturation (SpO2) of <90% during hospital stay for all patients or during a labor/delivery stay >2 days for pregnant patients or (2) manual chart review, as needed, performed by a physician investigator (BKA) and trained chart abstractors to verify the presence of severe COVID-19 symptoms.

### Exposures

The study focused on mRNA-1273, as it was conducted as part of a regulatory commitment from Moderna to multiple health authorities. The mRNA-1273 product used during the study period was the original monovalent vaccine. The exposures of interest were 3 doses (versus 2 doses and versus unvaccinated) or 4 doses (versus 3 doses and versus unvaccinated) of mRNA-1273. We included both 50-μg and 100-μg doses for third and fourth doses, as dosing information was not available for vaccines given outside of KPSC.

### Covariates

We identified potential confounders a priori based on the literature. Variables collected from EHRs before specimen collection included body mass index, smoking, Charlson comorbidity score, frailty index, chronic diseases, immunocompromised status, autoimmune conditions, healthcare visits (outpatient, virtual, ED, and inpatient), preventive care (other vaccinations, screenings, and wellness visits), history of SARS-CoV-2 infection, and history of SARS-CoV-2 molecular tests. Additional variables at date of specimen collection included pregnancy status, socioeconomic status (Medicaid and neighbourhood median household income), KPSC physician/employee status, medical center area, month of specimen collection, and specimen type (nasopharyngeal versus saliva).

### Statistical analyses

We described the distribution of SARS-CoV-2 variants by mRNA-1273 dose and calendar time. We compared the characteristics of cases and controls using the χ^2^ test or Fisher exact test for categorical variables and two-sample *t-*test or Wilcoxon rank-sum test for continuous variables, calculating the ASD to assess the balance of covariates. We used logistic regression adjusting for potential confounders to assess odds ratio (OR) and 95% CI for 3 doses versus unvaccinated or 4 doses versus unvaccinated of mRNA-1273 against infection and hospitalization with Omicron subvariants (BA.1 [not assessed for 4 doses], BA.2 [excluding BA.2.12.1], BA.2.12.1, BA.4, and BA.5 [BA.4/BA.5 were combined for hospitalization models]). We calculated VE (%) as (1 – OR) x 100 when OR was ≤1, and ([1/OR] – 1) × 100 when OR was >1. We also assessed 3-dose and 4-dose effectiveness against infection with Omicron subvariants by time since receipt of third or fourth dose of mRNA-1273 (for 3-dose VE: 14–30 days, 31–90 days, 91–150 days, and >150 days since the third dose; for 4-dose VE: 14–30 days, 31-90 days, and >90 days since the fourth dose).

To evaluate the incremental effectiveness of a) 3 doses versus 2 doses and b) 4 doses versus 3 doses of mRNA-1273, we further evaluated the rVE using the same approach as noted previously. Cases or controls receiving 2 doses or 3 doses of mRNA-1273, respectively, were combined as the comparison groups. The rVE by time since receipt of the third dose or the fourth dose against infection with Omicron subvariants was also assessed (for 3-dose versus 2-dose rVE: 14–30 days, 31–90 days, 91– 150 days, and >150 days since the third dose; for 4-dose versus 3-dose rVE: 14–30 days, 31-90 days, and >90 days since the fourth dose). rVE is interpreted as the incremental effectiveness of receiving an additional dose of mRNA-1273 compared with those who only received 2 doses or 3 doses, respectively.

Covariates included for adjustment across models were matching variables (age, sex, race/ethnicity, month of specimen collection) and other covariates with ASD>0.1 and *P*<0.1 from the comprehensive list of prespecified potential confounders. Additionally, we adjusted 3-dose versus 2-dose models for time between the second dose and specimen collection date and adjusted 4-dose versus 3-dose models for time between the third dose and specimen collection date to help account for possible differences in the timing of the second/third dose, respectively. SAS 9.4 software (SAS Institute) was used for all analyses.

We also conducted two sets of sensitivity analyses. In the first set of sensitivity analyses, we included cases that failed sequencing. For these analyses, according to the distribution of SARS-CoV-2 variants by month among successfully sequenced cases at KPSC, SGTF status and calendar month were used as a proxy to impute variant among cases that failed sequencing: specimens that were SGTF+ and collected during January-April 2022 were considered BA.1, those that were SGTF+ and collected during May-June 2022 were considered BA.4/BA.5 (combined, as it is not possible to distinguish BA.4 and BA.5 based on SGTF status), those that were SGTF- and were collected during January 2022 were considered Delta, and those that were SGTF- and were collected during February-June 2022 were considered BA.2. In the second set of sensitivity analyses, we excluded immunocompromised subjects to estimate the VE and rVE in the immunocompetent subjects. Because separate analyses with immunocompromised subjects only were not feasible given the small numbers across subvariants, both immunocompetent and immunosuppressed subjects were included in the main analyses.

## Data Availability

Individual-level data reported in this study are not publicly shared. Upon reasonable request and subject to review, KPSC may provide the de-identified aggregate-level data that support the findings of this study. De-identified data may be shared upon approval of an analysis proposal and a signed data access agreement.

## Acknowledgements

The authors would like to acknowledge the following Kaiser Permanente Southern California staff: Kourtney Kottman, Ana Acevedo, Elmer Ayala, Samantha Quinones, Samantha Baluyot, Jared Davis, Jose Rodriguez, Samuel Payan, Joanna Truong, Vanessa Pan, Katy Taylor, and Sarbijt Kaur-Chand for their technical and laboratory support in processing SARS-CoV-2 specimens; Julie Stern, Joy Gelfond, and Lee Childs for their coordination in processing SARS-CoV-2 specimens; and Radha Bathala, Maria Navarro, Elsa Olvera, Joy Gelfond, Jonathan Arguello, Kourtney Kottmann, Joanna Truong, and Diana Romero for their contributions to manual chart reviews of the electronic health records. The authors would like to acknowledge Helix OpCo, LLC, for their whole genome sequencing of SARS-CoV-2 specimens. The authors would also like to acknowledge the contributions by Moderna staff: Groves Dixon, PhD, and Julie Vanas. The authors thank the patients of Kaiser Permanente for their partnership with us to improve their health. Their information, collected through our electronic health record systems, leads to findings that help us improve care for our members and can be shared with the larger community.

## AUTHOR CONTRIBUTIONS

H.F.T, B.K.A, K.J.B, L.S.S, J.H.K, A.F., C.A.T and L.Q, were involved in the study concept and design. All authors were involved in acquisition, analysis, or interpretation of data. H.F.T, B.K.A, K.J.B and C.A.T drafted the manuscript. L.S.S, J.E.T, G.S.L, J.H.K, A.F, Y.L, S.Q, S.K.C, H.S.T, M.A, Y.D.P, S.C, L.Q, and C.A.T critically revised the manuscript for important intellectual content. J.E.T, Y.L, S.Q, and LQ conducted the statistical analyses. H.F.T, L.S.S, G.S.L, and S.C provided administrative, technical, or material support. H.F.T and S.C obtained funding and provided supervision.

## Ethics declarations

### Ethical approval

The study was reviewed and approved by the KPSC Institutional Review Board (IRB #12758). All study staff with access to protected health information were trained in procedures to protect the confidentiality of subject data. A waiver of informed consent was obtained as this is an observational study of authorized and recommended Moderna COVID-19 vaccine administered in the course of routine clinical care. To facilitate the conduct of this study, a waiver was obtained for written HIPAA authorization for research involving use of the EHR.

## Supplementary Information

### Supplementary Tables

Supplementary Table 1 Characteristics of SARS-CoV-2 specimens, by sequencing status and mRNA-1273 vaccination status

Supplementary Table 2a Vaccine effectiveness of 3 doses of mRNA-1273 against infection with SARS-CoV-2 variants by time since vaccination

Supplementary Table 2b Relative vaccine effectiveness of 3 versus 2 doses of mRNA-1273 against infection with SARS-CoV-2 variants by time since vaccination

Supplementary Table 3a Vaccine effectiveness of 4 doses of mRNA-1273 against infection with SARS-CoV-2 variants by time since vaccination

Supplementary Table 3b Relative vaccine effectiveness of 4 versus 3 doses of mRNA-1273 against infection with SARS-CoV-2 variants by time since vaccination

Supplementary Table 4a Vaccine effectiveness of 3 doses of mRNA-1273 against COVID-19 hospitalization associated with SARS-CoV-2 variants

Supplementary Table 4b Relative vaccine effectiveness of 3 versus 2 doses of mRNA-1273 against COVID-19 hospitalization associated with SARS-CoV-2 variants

Supplementary Table 5a Vaccine effectiveness of 4 doses of mRNA-1273 against COVID-19 hospitalization associated with SARS-CoV-2 variants

Supplementary Table 5b Relative vaccine effectiveness of 4 versus 3 doses of mRNA-1273 against COVID-19 hospitalization associated with SARS-CoV-2 variants

Supplementary Table 6a Vaccine effectiveness of 3 doses of mRNA-1273 against infection with SARS-CoV-2 variants by time since vaccination, using SGTF data to impute unidentified subvariants

Supplementary Table 6b Relative vaccine effectiveness of 3 versus 2 doses of mRNA-1273 against infection with SARS-CoV-2 variants by time since vaccination, using SGTF data to impute unidentified subvariants

Supplementary Table 7a Vaccine effectiveness of 4 doses of mRNA-1273 against infection with SARS-CoV-2 variants by time since vaccination, using SGTF data to impute unidentified subvariants

Supplementary Table 7b Relative vaccine effectiveness of 4 versus 3 doses of mRNA-1273 against infection with SARS-CoV-2 variants by time since vaccination, using SGTF data to impute unidentified subvariants

Supplementary Table 8a Vaccine effectiveness of 3 doses of mRNA-1273 against COVID-19 hospitalization associated with SARS-CoV-2 variants, using SGTF data to impute unidentified subvariants

Supplementary Table 8b Relative vaccine effectiveness of 3 versus 2 doses of mRNA-1273 against COVID-19 hospitalization associated with SARS-CoV-2 variants, using SGTF data to impute unidentified subvariants

Supplementary Table 9a Vaccine effectiveness of 4 doses of mRNA-1273 against COVID-19 hospitalization associated with SARS-CoV-2 variants, using SGTF data to impute unidentified subvariants

Supplementary Table 9b Relative vaccine effectiveness of 4 versus 3 doses of mRNA-1273 against COVID-19 hospitalization associated with SARS-CoV-2 variants, using SGTF data to impute unidentified subvariants

Supplementary Table 10a Vaccine effectiveness of 3 doses of mRNA-1273 against infection with SARS-CoV-2 variants by time since vaccination, excluding immunocompromised patients

Supplementary Table 10b Relative vaccine effectiveness of 3 versus 2 doses of mRNA-1273 against infection with SARS-CoV-2 variants by time since vaccination, excluding immunocompromised patients

Supplementary Table 11a Vaccine effectiveness of 4 doses of mRNA-1273 against infection with SARS-CoV-2 variants by time since vaccination, excluding immunocompromised patients

Supplementary Table 11b Relative vaccine effectiveness of 4 versus 3 doses of mRNA-1273 against infection with SARS-CoV-2 variants by time since vaccination, excluding immunocompromised patients

Supplementary Table 12a Vaccine effectiveness of 3 doses of mRNA-1273 against COVID-19 hospitalization associated with SARS-CoV-2 variants, excluding immunocompromised patients

Supplementary Table 12b Relative vaccine effectiveness of 3 versus 2 doses of mRNA-1273 against COVID-19 hospitalization associated with SARS-CoV-2 variants, excluding immunocompromised patients

Supplementary Table 13a Vaccine effectiveness of 4 doses of mRNA-1273 against COVID-19 hospitalization associated with SARS-CoV-2 variants, excluding immunocompromised patients

Supplementary Table 13b Relative vaccine effectiveness of 4 versus 3 doses of mRNA-1273 against COVID-19 hospitalization associated with SARS-CoV-2 variants, excluding immunocompromised patients

### Supplementary Figures

Supplementary Fig. 1: Distribution of SARS-CoV-2 variants by mRNA-1273 vaccination status

Supplementary Fig. 2: Distribution of SARS-CoV-2 variants by month of specimen collection

